# Modelling digital and manual contact tracing for COVID-19 Are low uptakes and missed contacts deal-breakers?

**DOI:** 10.1101/2021.04.29.21256307

**Authors:** Andrei Rusu, Katayoun Farrahi, Rémi Emonet

## Abstract

Comprehensive testing schemes, followed by adequate contact tracing and isolation, represent the best public health interventions we can employ to reduce the impact of an ongoing epidemic when no or limited vaccines are available and the implications of a full lockdown are to be avoided. However, the process of tracing can prove feckless for highly-contagious viruses such as SARS-Cov-2. The interview-based approaches often miss contacts and involve significant delays, while digital solutions can suffer from insufficient adoption rates or inadequate usage patterns. Here we present a novel way of modelling different contact tracing strategies using a generalized *multi-site mean-field* model, which can naturally assess the impact of both manual and digital approaches. Our methodology can readily be applied to any compartmental formulation, thus enabling the study of several complex pathogens. We use this technique to simulate a new epidemiological model, SEIR-T, and show that, given the right conditions, tracing in a COVID-19 epidemic can be effective even when digital uptakes are sub-optimal or interviewers miss a fair proportion of the contacts.

## 1 INTRODUCTION

The epidemic started in Wuhan, China by the SARS-Cov-2 virus has rapidly spread in communities from all around the world, rapidly becoming a major global threat which, as of February 2021, is responsible for more than 100 million infected people and two million deaths ^1^. Prompted by the scale of this disease, cross-disciplinary teams started working against the clock to develop reliable pathogen spreading models that could be used to assess the effectiveness of different public health interventions. Since imposing a general lockdown has proven economically unbearable for most countries, the attention significantly shifted to less restrictive yet partially successful measures, such as educating the public to socially distance, deploying large-scale testing and quarantining contacts through various tracing mechanisms (Dighe et al., 2020). The latter proved rather challenging for the traditional interview-based approaches, mostly due to staffing issues and a generally poor recollection exhibited by the interviewees. As a result, digital alternatives were quickly sought after by several governments. These were or are currently being deployed in many states, most being reliant on either a Bluetooth solution, such as the Exposure Notification (GAEN) system (Google and Apple, 2020), or a geolocation-based software, similar to the Integrated Disease Surveillance Programme in India (Garg et al., 2020). That being said, the efficiency of these strategies remains largely dependent on the application adoption rates and the behavioral patterns of their userbase (i.e. self-isolation compliance, respecting the usage guidance, keeping the tracing device turned on etc.). Although some have suggested an application uptake of at least 50% would be needed at the population level to contain the epidemic (Ferretti, 2020), while others showed via simulations that 60% would be enough to stop the spread without requiring further interventions (Robert Hinch et al., 2020), the adoption levels generally quoted in the literature as “sufficient” remain mostly unattainable due to privacy concerns and internet access limitations. The picture gets even more intricate when the aforementioned behavioral issues are widespread in the active users’ communities or if inadequate testing regimes and manual tracing procedures are employed.

Motivated by the limited evidence we have on the efficacy of contact tracing methods in the face of such challenges, we developed a *multi-site mean-field* model that can simulate the joint effects of these variables on the evolution of an epidemic, and used it to study COVID-19 via a new disease-specific compartmental formulation - SEIR-T, see Fig 1. Our methodology draws inspiration from the work of Farrahi et al. (2014), but it enables the simulation of more varied scenarios involving digital tracing at different uptake levels *r*, manual tracing with various network overlaps Γ, or both procedures combined. Moreover, we propose separating the “traced” status from the infection states, thus allowing for a node to get isolated at all times (unless it has reached an end state, i.e. recovered or dead), ensuring self-isolation can end due to non-compliance or term expiration, and halting any impact it might have had on an individual’s normal disease progression. This feature also makes our approach directly compatible with any compartmental model. As is customary, all our code was made publicly available (see appendix A.1).

**Figure 1:**
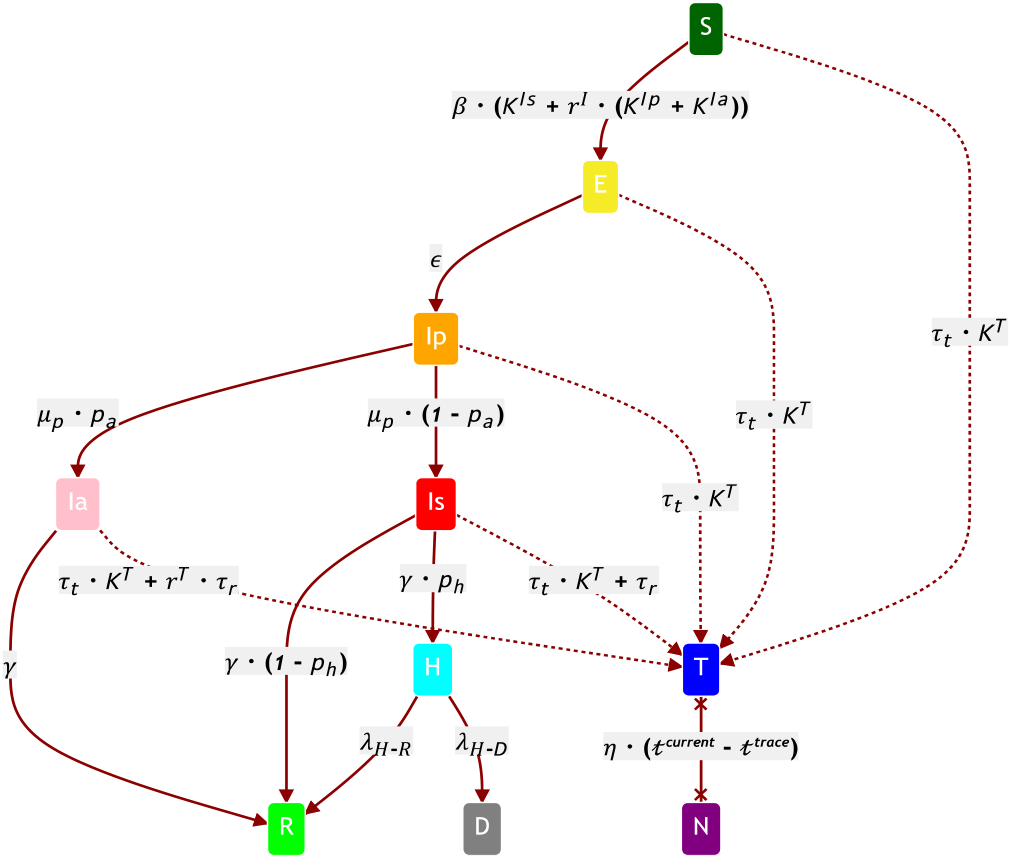
The SEIR-T Compartmental Model for COVID-19. Each node has 2 allocated variables: an infection state and a tracing status. The infection states from top to bottom are: *S* - susceptible; *E* - exposed but not infectious; *I*_*p*_ - infectious, presymptomatic; *I*_*a*_ - infectious, asymptomatic; *I*_*s*_ - infectious, symptomatic; *H* - hospitalized; *R* - recovered / removed; *D* - dead. At any point in time, a node’s tracing status can either be *T* (traced and isolated) or *N* (not traced/isolated or non-compliant). Each state transition has a certain time-dependent probability *p*_*S*1 → *S*2_; the edge labels here represent both 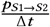, and the *λ* rate of the corresponding exponential to sample from in CT simulations.

The experiments we conducted confirm that the potency of contact tracing not only depends on the accuracy of the tracing network, but also on several other variables (i.e. testing rates, tracing reliability, staffing and delays, public-health communiqués, isolation conformity etc.), an optimal configuration of these given a country’s epidemiological situation being essential for a swift viral containment. Even when low uptakes are registered (*r* ≤ 0.4)or the interviewing process misses many contacts (Γ ≤ 0.5), our simulations suggest that significant reductions in the peak of infections and the total number of deaths can still be achieved given small tracing delays and the appropriate levels of testing and self-isolation compliance. What is more, the combined effects of manual and digital tracing can drive the effective reproduction number *R* below 1 even when neither is very efficient. We validate our results on different hyperparameter combinations and network topologies, including random Erdős–Rényi (Erdös and Rényi, 1959), scale-free, and small-world graphs (Xiao Fan Wang and Guanrong Chen, 21).

## 2 RELATED WORK

In recent years, modeling epidemics has mainly been achieved via either of two paradigms: agent-based (ABM) or equation-based models (EBM). The first represents a bottom-up approach in which a set of behaviors are attributed to each agent in a topological system. These behaviors dictate every individual’s action patterns through this topology, and ultimately determine the execution of different *discrete* interaction events (e.g. infection spreading, tracing notification broadcasting etc.). ABMs tend to be relatively complex and resource-intensive to simulate, the involved cost being often justified by their exhibited level of granularity and their ability to monitor public interventions at the individual level (Sukumar and Nutaro, 2012). Government-advising groups in the UK decided to employ this paradigm early on in the COVID-19 pandemic to estimate the effects of such interventions (Ferguson et al., 2020; Hinch et al., 2020). A more recent Oxford study looked at the combined effects of manual tracing with digital solutions, at various application uptakes, via a rich yet scalable ABM fitted to mobility data from different counties in Washington (Abueg et al., 2020). We consider their findings the strongest modeling evidence to date that digital tracing can be effective even at low adoption rates.

On the other hand, EBMs define a set of equations that express the evolution of certain *continuous* observables over time. These generally represent system states (called compartments) showing how a disease progresses through a population. SIR, a widely-known EBM, utilizes three ordinary differential equations to model a generic epidemic (Kermack et al., 1927). Extensions of SIR were subsequently used to simulate the transmission of many pathogens, including Zika (Aik et al., 2017), Ebola (Berge et al., 2017), and most recently SARS-Cov-2 - e.g. SIDARTHE (Giordano et al., 2020), SUQC (Zhao and Chen, 2020). The present study employs a variation of the compartmental model designed by the French National Institute of Health and Medical Research to study the impact of lockdown exit strategies on the COVID-19 spread (Di Domenico et al., 2020).

At the intersection of these two paradigms lies the category of *multi-site mean-field* models which combine the mathematical rigour and the superior generalizability of EBMs with the ability to leverage locality information regarding every individual. Similarly to ABMs, the infection spreads over a predefined network that can either be random (Rozhnova and Nunes, 2009) or inferred from real data (Farrahi et al., 2014), yet unlike ABMs, the dynamics are fully characterized by state transition equations. Tsimring and Huerta (2003) first employed this technique for modeling contact tracing in a generic epidemic. Farrahi et al. (2014) took this idea one step further by restricting the tracing propagation to a subset of the infection network, thus accounting for the inherently noisy nature of this process. Even though both of these exhibited powerful modeling capabilities, they were limited by their underlying “SIRT” formulation which made several unrealistic assumptions that do not generalize to real viral diseases: inter alia, the recovery was conditioned on tracing, susceptibles could not be wrongfully isolated, a traced person remained noninfectious for the full duration of the epidemic. Our model fixes these issues by separating the traced/isolated status from the infection state, therefore allowing for all the “active” nodes (i.e. not hospitalized, recovered or dead) to become traced or exit self-isolation after a certain time without changing their corresponding disease progression (see Fig 1). Concurrently, this modification enables one to simulate the effects of contact tracing independently of the compartmental formulation utilized.

For the sake of completeness, we would also like to mention that branching process models for epidemics have become increasingly popular in the last few years (Jacob, 2010; Lashari and Trapman, 2018). One such model, concerned with studying the effects of manual contact identification together with digital tracing solutions at various uptakes on the COVID-19 pandemic, has recently been proposed (Plank et al., 2020). Its simulations show that effective manual tracing needs to be coupled with an application uptake of at least 75% to achieve containment, although smaller adoption rates can decrease the reproduction number *R* if combined with other public health interventions. Our results are in accordance with the latter observation, but they also show that, given the right testing and tracing regimes, lower and achievable adoption levels are enough to significantly reduce the viral spread.

## 3 MODEL OUTLINE

Motivated by growing evidence that simple SIR frameworks are inefficient at capturing the dynamics of SARS-Cov-2 epidemics (Moein et al., 2021), we propose a new compartmental model (Fig 1) that accounts for many of its particular features. A description of the parameters involved in our simulations, together with the values we consider for each of them, can be consulted in Table 1.

**Table 1:** Compartmental model parameters used in our simulations.

| Parameter | Value(s) | Description |
| --- | --- | --- |
| $\beta$ | .0791 | Transmission rate corresponding to $R_0 = 3.18$.<br>According to inference by Di Domenico et al. (2020) |
| $K^{S_i}$ | $\in \mathbb{N}$ | Number of neighbors in state $S_i$ in a given network |
| $r^I$ | 0.5 / 1 | Relative infectiousness of $I_p$ and $I_a$ compared to $I_s$.<br>Source for 0.5: Grassly et al. (2020).<br>But weak evidence as per Mc Evoy et al. (2020) |
| $\epsilon^{-1}$ | 3.7 (days) | Latency period. Source: Di Domenico et al. (2020) |
| $p_a$ | 0.2 / 0.5 | Probability of being asymptomatic. Still disputed:<br>- 0.2: Di Domenico et al. (2020); Mizumoto et al. (2020)<br>- 0.5: Keller et al. (2020); Oran and Topol (2020) |
| $\mu_p^{-1}$ | 1.5 (days) | Presymptomatic period. Source: Ferretti et al. (2020) |
| $p_h$ | .1 | Probability of being hospitalized for *adults*.<br>Equivalent to $p_{ss}$ in Di Domenico et al. (2020) |
| $\gamma^{-1}$ | 2.3 (days) | Infectious period with mean generation time 6.6 days.<br>Source: Di Domenico et al. (2020) |
| $\lambda_{H-R}$ | .083 | Daily rate of recovery for *adults*. Source: Etalab (2020) |
| $\lambda_{H-D}$ | .0031 | Daily rate of death for *adults*. Source: Etalab (2020) |
| $\tau_t$ | [0-0.5] | Contact tracing rate. It encompasses many related phenomena:<br>Tracing latency/efficiency due to staffing/server reliability,<br>Likelihood of being isolated given no. of traced neighbors.<br>Can range from no tracing to every 2 days on average |
| $\tau_r$ | [0-0.5] | Testing / Random tracing rate.<br>Can range from no testing to every 2 days on average |
| $r^T$ | .8 | Relative probability for $I_a$ to be tested positive (against $I_s$).<br>We assume $E$ and $I_p$ testing mostly results in false negatives |
| $\eta$ | 0 / .001 | Non-compliance / Self-isolation exit rate.<br>Scaled by the time elapsed since isolating: $t^{\text{current}} - t^{\text{trace}}$ |

Our propagation model consists of a predefined network onto which the infection spreads, and either one (*dual*) or two (*triad*) subnetworks through which one type of contact tracing gets conducted (manual, digital, or both for triads). Connected vertices in the first network are to be considered “close contacts”, as defined by institutions like the CDC ^2^. The other graphs are generally a subset of the first, missing edges being equivalent to misuse in the case of app-based tracing or contacts not recalled in the manual interview processes, while nodes with no links are to represent individuals that never use the application. We control both of these aspects via two interlinked hyperparameters: *degree of overlap* 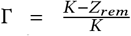, where *K* is the infection network’s mean degree, from which an average of *Z*_*rem*_ edges are randomly removed to obtain a tracing graph; and *uptake rate r* that controls the *maximum percentage* of nodes with at least one link in a tracing subnetwork. All our experiments focus on the impact of varying either of them. We assume a ‘traced’ (*T* state) individual automatically enters self-isolation, so infecting or getting infected remains impossible until it becomes ‘non-isolating’ (*N*). Moreover, we presume that a node’s probability to get infected increases with the number of infectious neighbors it has, while the likelihood of being traced and compliant with isolation scales up with the total number of adjacent *T* nodes in each tracing network.

## 4 SIMULATIONS OVERVIEW

Most simulations in this study were run over Erdős–Rényi random graphs with different population sizes and average degrees. It is worth mentioning that this type of graph model may not correctly capture the interaction patterns of certain social networks (Newman et al., 2002), but it tends to offer acceptable approximations most of the time. That being said, experiments involving scale-free (SF) and small-world (SW) networks are briefly covered in Section 5.4, while we note that users’ mobility data or ABM simulations can be used to bias the link distributions further. As SARS-Cov-2 is an overdispersed pathogen (Adam et al., 2020; Endo et al., 2020), SFs should offer a more accurate representation of the transmission chain since super-spreaders can be factored in by contact graphs whose degrees are distributed according to power-laws. On the other hand, SWs more closely resemble interactions in social networks due to their larger clustering coefficient, while clusters being known as important drivers of this pandemic (Liu et al., 2020).

In our model the time intervals between two state changes of the same kind are assumed to form an exponential distribution, with the *λ* rate equal to the corresponding transition label of Fig 1. For efficiency, we simulate the COVID-19 epidemic using Gillespie’s algorithm (Gillespie, 1977), which has been shown to be *stochastically exact* and faster than the Monte Carlo (MC) method for both static and dynamic network-based applications (Vestergaard and Génois, 2015). Compared to a continuous-time MC simulation, which entails sampling the next transition for all the possible state changes, discarding all but the most “recent” event (Farrahi et al., 2015), Gillespie’s procedure directly draws the time elapsed until the next transition and identifies the state change most likely to have taken place in that period (see Algorithm 1).

## 5 RESULTS AND DISCUSSION

### 5.1 Variation induced by population size

Early simulations suggested that the degree of variability across runs scales with the number of participant nodes. In order to verify this hypothesis, we design an experiment in which we vary the population size: *N* ∈ {200, 500, 1000, 2000, 5000, 10000, 20000}, while keeping the other hyperparameters fixed at: *K* = 10, uptake *r* = 0.5 (Γ implicitly derived), *p*_*a*_ = 0.2, *τ*_*t*_ = 0.1, *τ*_*r*_ = 0.1, with one infectious individual set for time *t*_*0*_. The statistics in Fig 2 represent averages over several simulations conducted with 10 different network initializations, filtering out the iterations with less than three infected (for a total of 80-100 simulations per each *N*). The data confirm the variance in peaks of infection increases as the network expands, aspect which can be explained by the growing difference between early-stopped and full-blown outbreaks. In contrast, the uncertainty in estimating the relative percentage of these maximal points expectedly decreases with the size, a choice of *N* = 1000 resulting in a tolerable variance of almost 3%, while *N* = 10000 ensures an even smaller variability of <1% across runs. Consequently, we consider these two values representative for our model’s expressive power and use them both in our experiments, with the mention that we account for the difference in variances by simulating 7 different networks with 15 random seeds each for *N* = 10000, but 50 networks and 15 seeds in the case of *N* = 1000.

**Figure 2:**
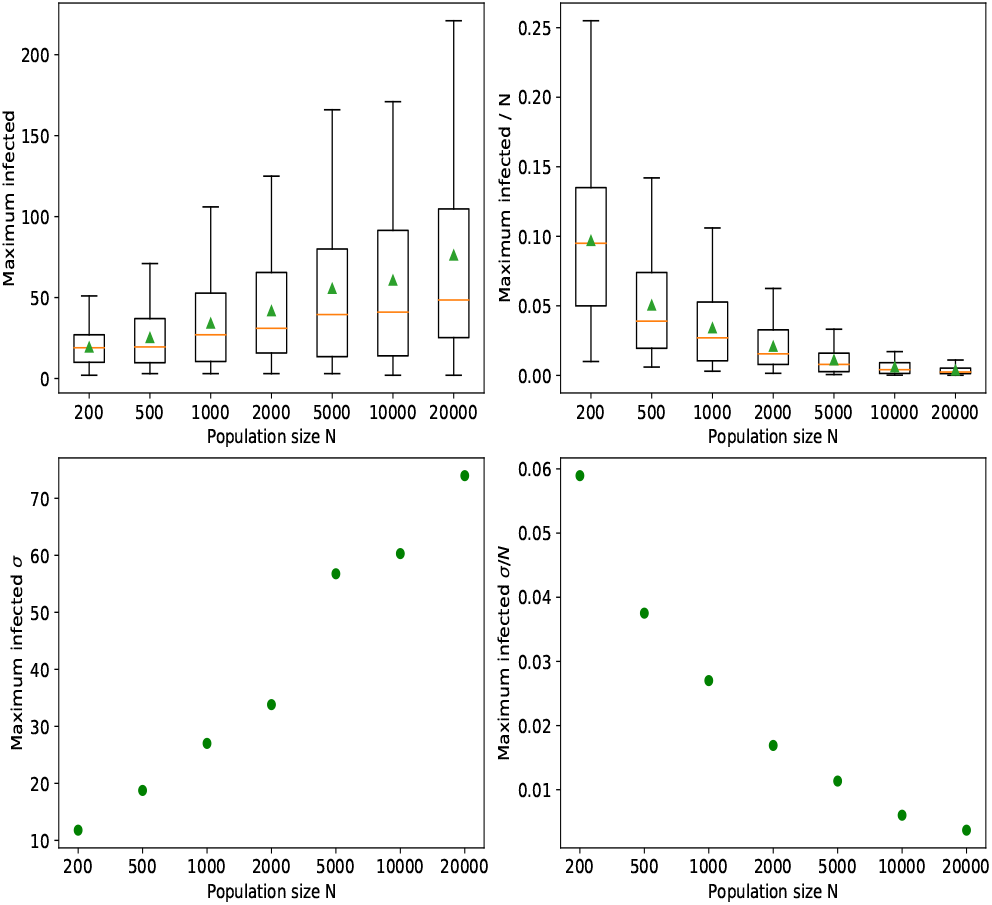
Uncertainty of simulation results with respect to the infection peak. Values from 80-100 runs plotted for different population sizes (boxplots on top, standard deviations below). The left side displays absolute values, whereas on the right numbers are scaled down by *N*.

### 5.2 Tracing overlap in larger populations

Next, we want to assess the impact of varying the tracing network’s accuracy in an outbreak involving a large community of *N* = 10000 individuals. To achieve this, we use the following hyperparameters: Γ ∈ {0.1, 0.2, …, 1} (uptake *r* is implicitly derived), *K* = 10, *p*_*a*_ = 0.2, *τ*_*r*_ ∈ {0.001, 0.01, 0.04, 0.07, 0.1}, *τ*_*t*_ ∈ {0.01, 0.04, 0.07, 0.1}, *η* = 0 (assuming everybody self-isolates until they are no longer infectious), and a single *I*_*p*_ node at time *t*_*0*_. The figures below represent results averaged over 105 runs, as previously described.

Fig 3 shows that a sub-optimal test rate, such as *τ*_*r*_ = 0.001, leads to inconclusive results, where the variance induced by the stochasticity of the process shadows any benefit obtained through contact tracing. With better testing, clearer patterns start to emerge: The higher the contact tracing rate, the better the peak suppression is and the faster it gets approached (Fig 4). As *τ*_*r*_ becomes even more effective, smaller tracing network overlaps are needed to swiftly reduce that maximum point. Moderate contact tracing (*τ*_*t*_ ∈ {.04, .07}) achieves a delay in the peak for smaller Γ, and noticeable reductions for Γ ≥ 0.5. In contrast, *τ*_*t*_ = 0.01 seems too small to produce any positive effect. In real life, the latter scenario would occur if the tracing programme is slow and it misses many contacts or if the digital contacts application fails to promptly notify many of its active users.

**Figure 3:**
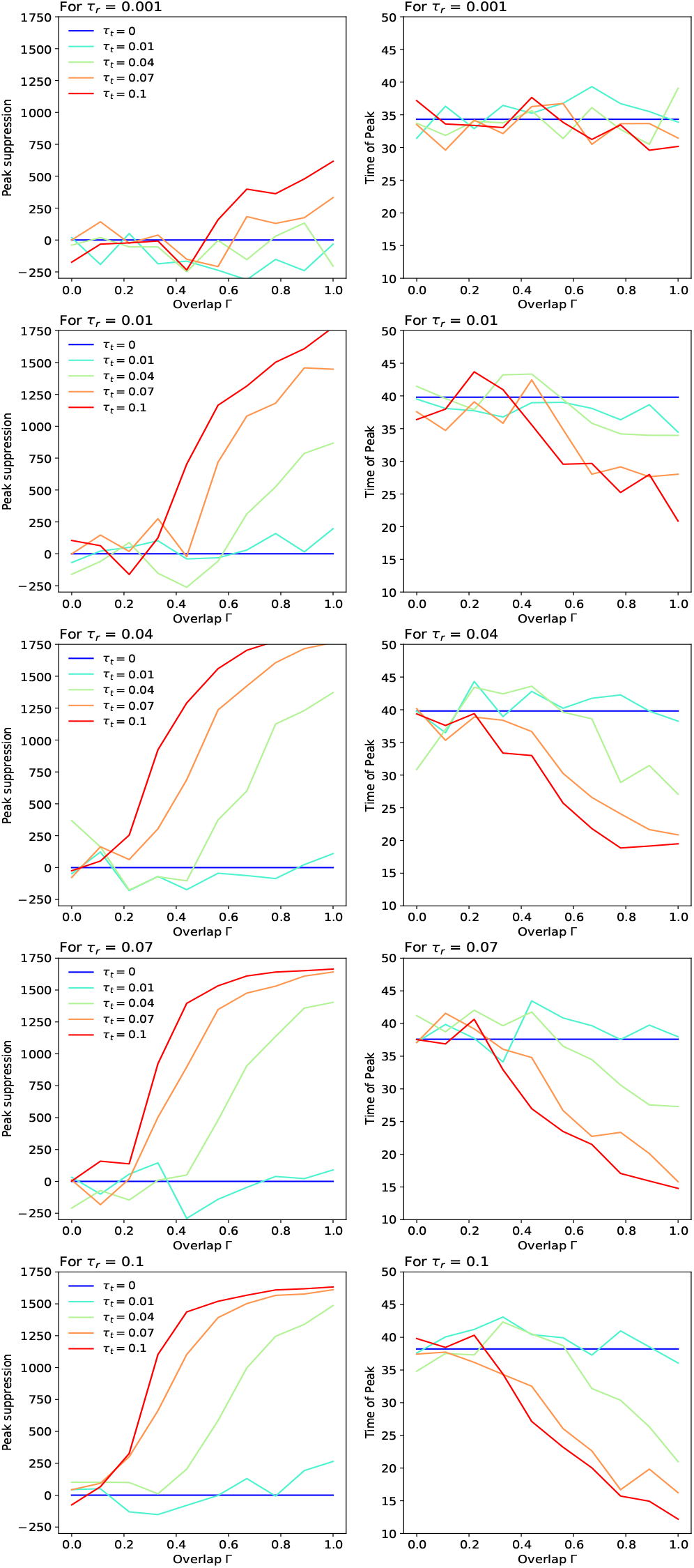
Peak suppression (left) and the time of peak (right) at various tracing network overlaps. Values are averaged over 105 runs. The suppression is calculated by subtracting the average maximal infected point given by each parameter configuration from the average point obtained with no contact tracing (*τ*_*t*_ = 0). Apart from *τ*_*r*_ = 0.001 and *τ*_*t*_ = 0.01 which produce inconclusive results, the effectiveness of a containment strategy expectedly scales with the testing and tracing rates.

**Figure 4:**
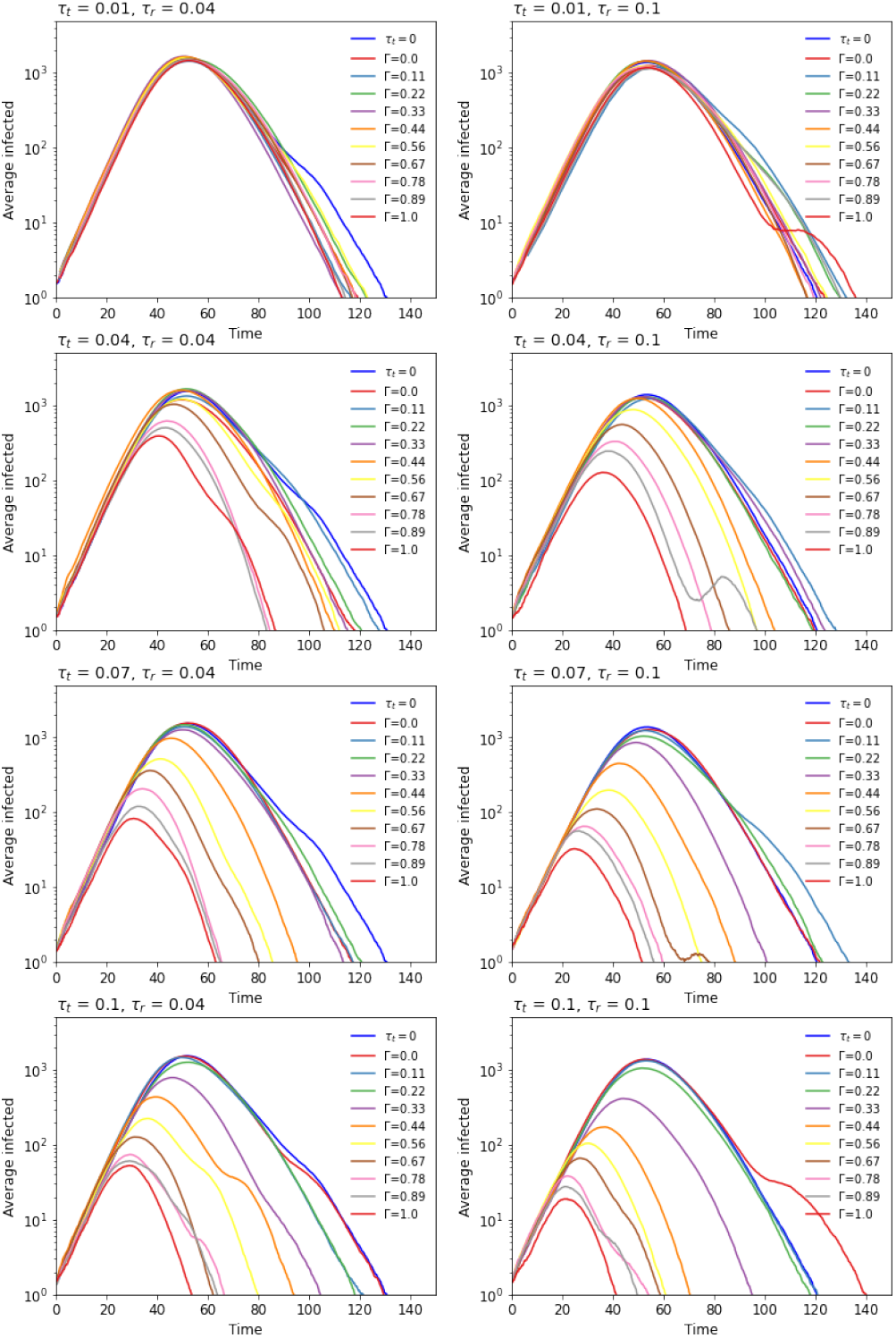
Epidemic evolution over time given a less efficient (left) and a more effective (right) testing regimes. Results averaged over 105 simulations. As the contact tracing rate increases, the accuracy of the network given by Γ becomes more important for “flattening” the curves. The case with no contact tracing (*τ*_*t*_ = 0) is colored in blue.

Aside from outlining the effects of the tracing network overlap and of different testing strategies, this experiment also reveals which parts of *τ*_*r*_ and *τ*_*t*_’s parameter space are more interesting to explore. As a result, we observe that meaningful results are obtained when both are ≥ 0.04, while values ≥ 0.1 fall within the “effective” region.

#### Algorithm 1

Event sampling via an adapted Gillespie algorithm

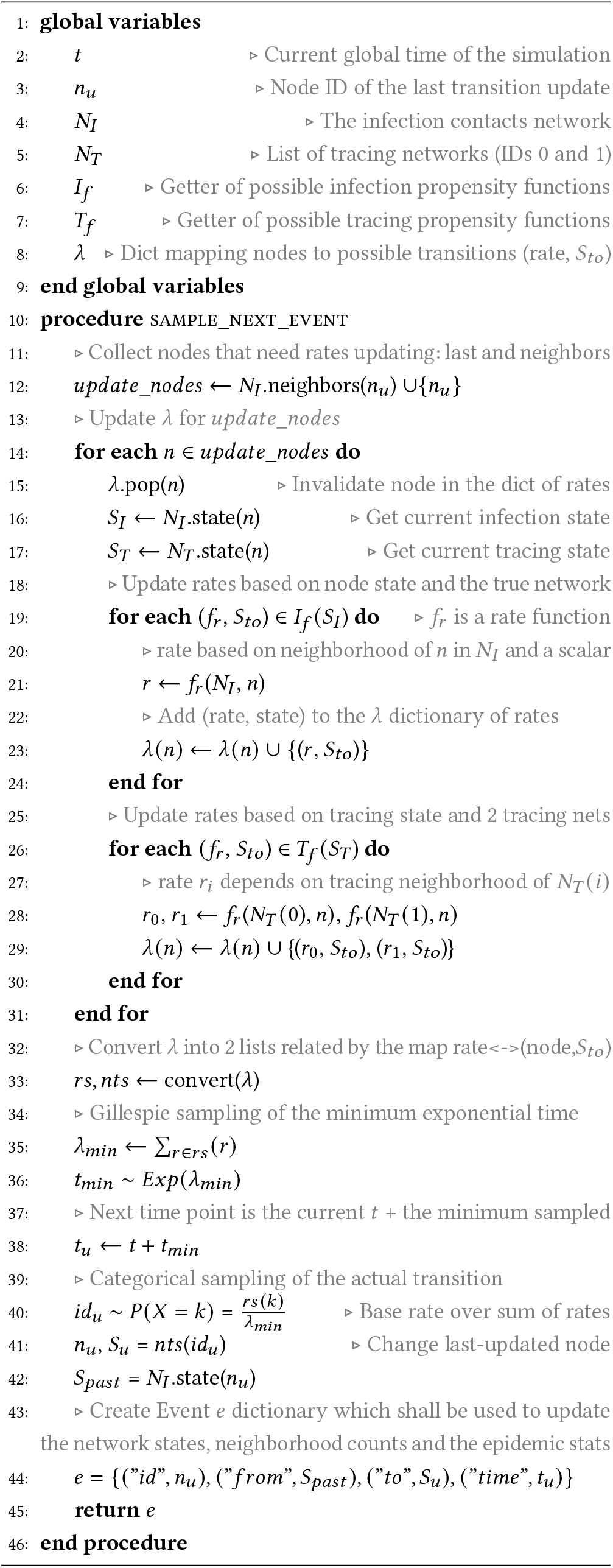

### 5.3 Effects of average degree and app uptake

Further, we explore the impact of application uptake in scenarios with different average degrees (*K* ∈ 10, 20) and more suitable “test and trace” strategies (*τ*_*t*_, *τ*_*r*_ ∈ {0.05, 0.1, 0.2, 0.5}). For this trial, we set *N* = 1000, *p*_*a*_ = 0.2, *η* = .001 (with automatic isolation exit after 14 days), and select a single *I*_*p*_ node as the infection seed. The results are averaged over 750 simulations to reduce the variance induced by the smaller population size.

Fig 5 shows the peak suppression achieved by each strategy given a specific adoption level. For *τ*_*t*_ = 0.05, uptakes *r* ≤ 0.5 generally give results within the noise region. Improving the contact tracing rate, however, leads to a noticeable decrease of this maximal point, even at smaller adoption levels. This is particularly true in the larger average degree case. Interestingly, deploying a wider-scale testing programme alone (*τ*_*r*_ = 0.5) seems to lead to a considerable spread reduction which makes contact tracing less beneficial at achievable uptakes (even entirely profitless in the *K* = 10 situation).

**Figure 5:**
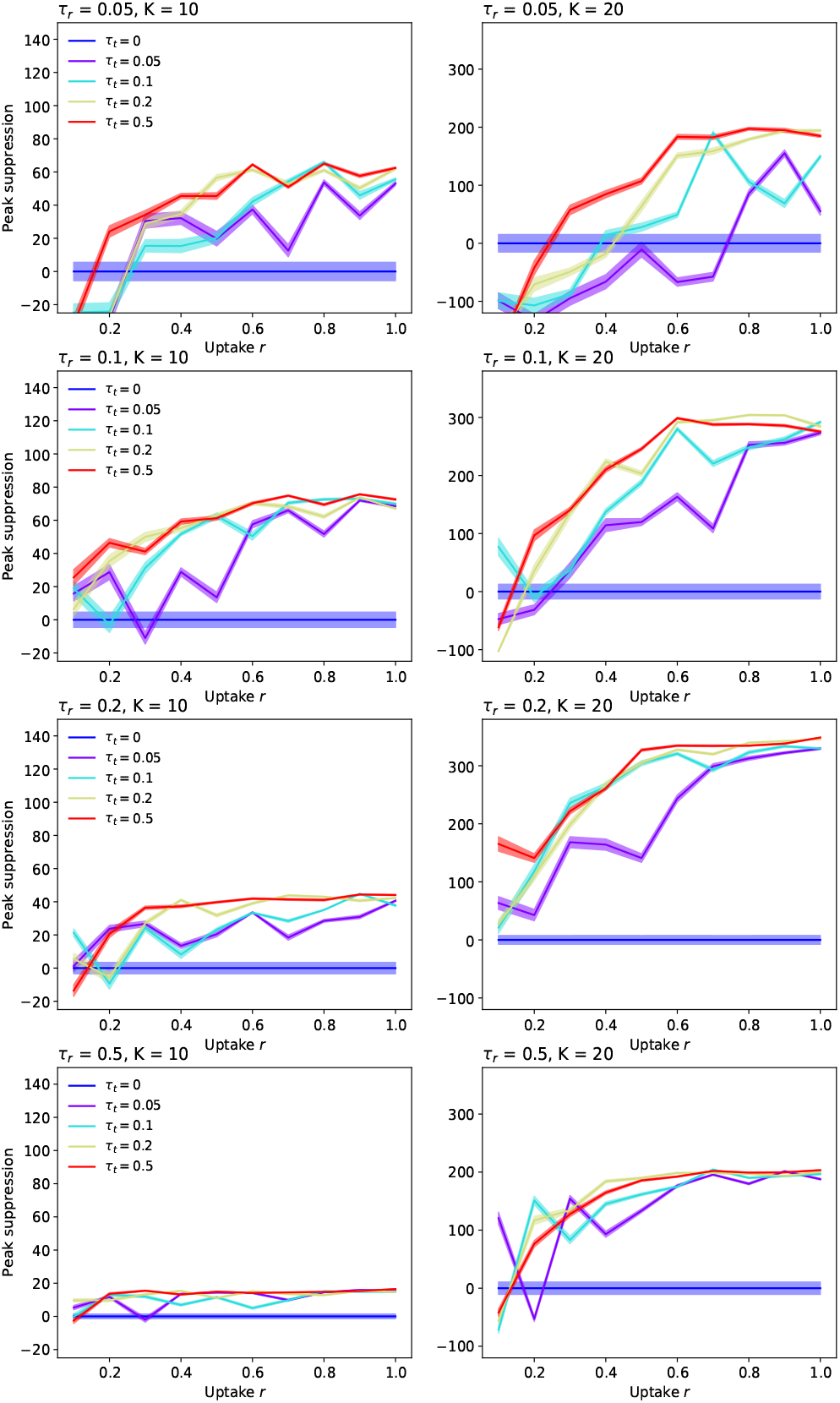
Uptake rate *r* against peak suppression. Suppression is difference in peak to no tracing, i.e. *τ*_*t*_ = 0. *K* = 10 given on the left, *K* = 20 on the right. The case with no contact tracing (*τ*_*t*_ = 0) is colored in blue. All lines were plotted with the 95% confidence intervals resulted from 750 different runs.

Our findings suggest that a testing rate of *τ*_*r*_ = 0.1 remains suitable in conjunction with contacts isolation not only for the previous experiment with *N* = 10000, but also in these smaller scale scenarios featuring different average degrees. Consequently, we decided to examine further the effect of such a testing regime on the evolution of the spread (Fig 6) and the number of total deaths (Fig 7) for *K* = 20. The first chart below illustrates how the epidemic curves significantly “flatten” for uptakes *r* ≥ 0.4, the effect being more apparent as the contact tracing rate increases. The second diagram puts these results into perspective by showing that the number of deaths can be reduced even with lower uptakes, while at the other end of the spectrum many simulations ended with notably fewer deceased (even none for effective tracing *τ*_*t*_ ≥ 0.2).

**Figure 6:**
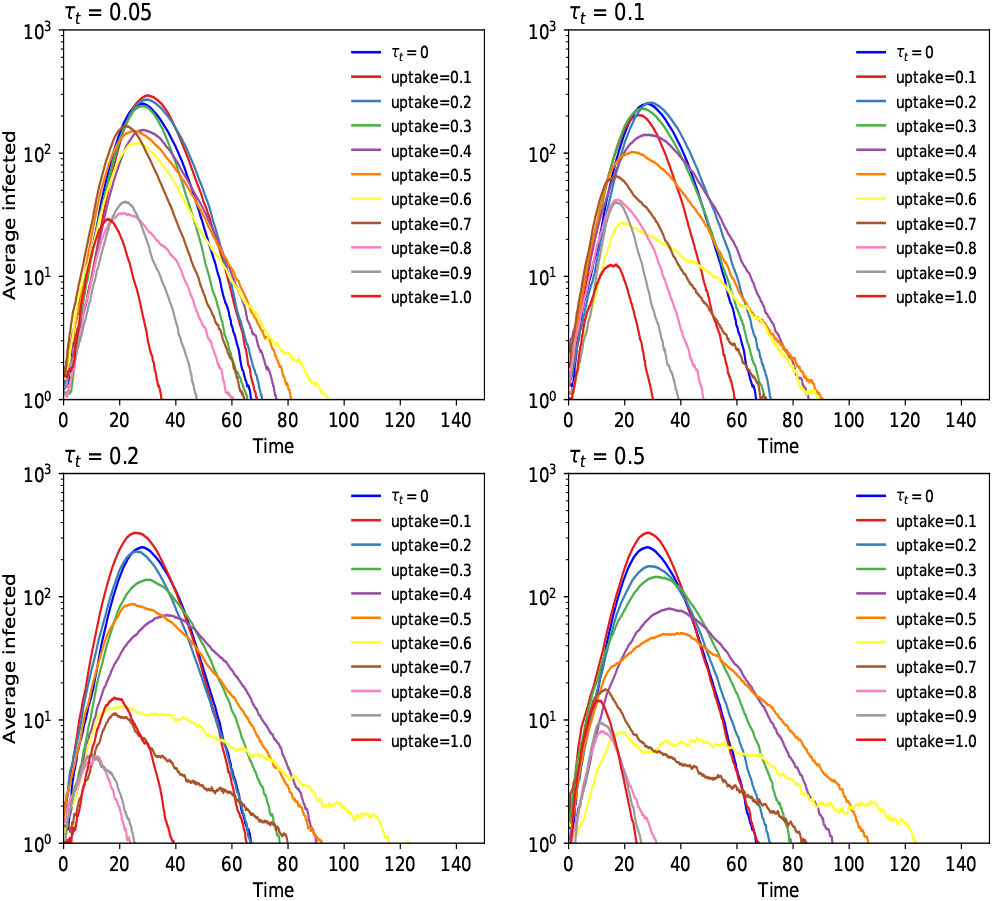
Epidemic evolution over time for *τ*_*r*_ = 0.1 and *K* = 20. Results here are for a population of *N* = 1000, and were averaged over 750 runs. The case with no tracing (*τ*_*t*_ = 0) is colored in blue.

**Figure 7:**
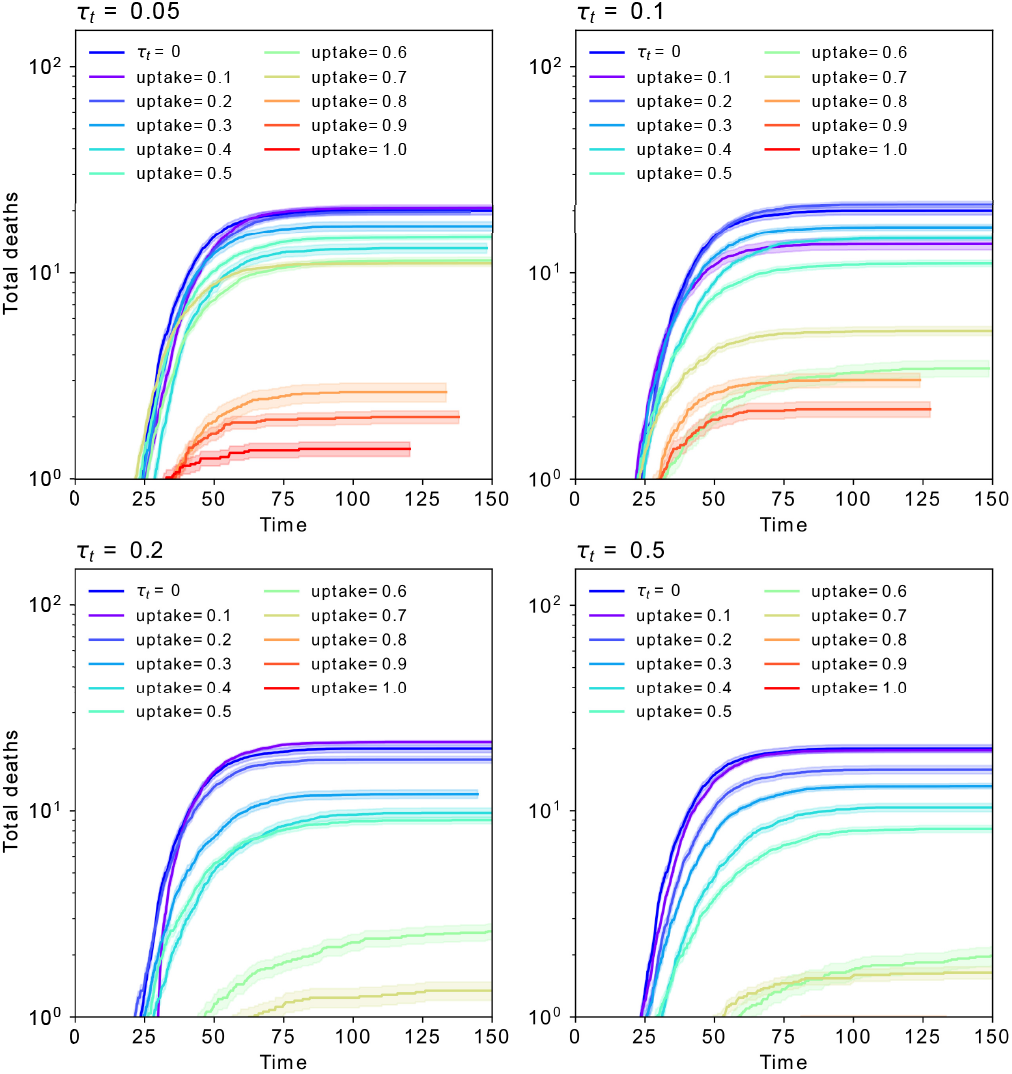
Total deaths over time for *τ*_*r*_ = 0.1 and *K* = 20. The 95% confidence intervals resulted from 750 runs are displayed around each line. The case with no contact tracing (*τ*_*t*_ = 0) is colored in blue.

### 5.4 Combining digital tracing with an imperfect manual tracing process

Finally, we look into a more realistic scenario in which digital solutions complement an inherently imperfect interview-based tracing system. To that end, we employ a *triad network* topology (example in Fig 8), with digital tracing happening at a rate of 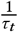 days on average over one subgraph given by the uptake *r* ∈ {0.1, 0.25, 0.4, 0.55, 0.7, 0.85, 1}, while the manual process gets carried at a slower pace of 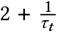 days on average over a third network whose edges are randomly removed according to overlap Γ ∈ {0.1, 0.25, 0.4, 0.55, 0.7, 0.85, 1}. For the purpose of this experiment, we use a more representative graph structure for the SARS-Cov-2 transmission based on the Holme and Kim (2002) model, which features both a SF degree distribution and a SW clustering coefficient. The network hyperparameters chosen here are: *N* = 1000, *m* = 10 (number of random edges to add for each new node) and *p* = 0.2 (probability of making a triangle after adding a random edge). To avoid runs in which the epidemic gets quickly contained by chance, we seed the simulation with 10% of *I*_*p*_ nodes. The other parameters remain unchanged from last section.

**Figure 8:**
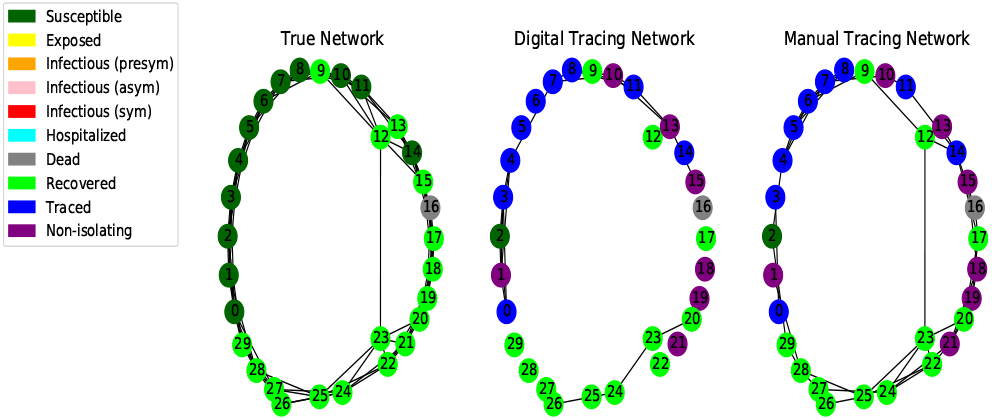
Final state of an epidemic simulation over a *triad* topology. Infection spreads with respect to the neighborhoods of the first network (here a SW graph); the second network corresponds to digital tracing at uptake *r* = 0.5, while the third involves manual tracing with overlap Γ = 0.5.

First aspect to notice in Fig 9 and Fig 10 is that all curves remain monotonic with respect to *r*, while the dissimilarities between different *τ*_*t*_ rates become more apparent than what was observable in the last experiment. This is a direct consequence of the increased number of infected selected for time *t*_*0*_, which prevents simulations from averaging over too many early-stopped runs. Considering the scale this pandemic has reached and the unavoidable presence of a delay between the infection onset and the debut of tracing, scenarios such as this one are more likely to occur, and therefore of a greater importance (Hellewell et al., 2020; Hinch et al., 2020).

**Figure 9:**
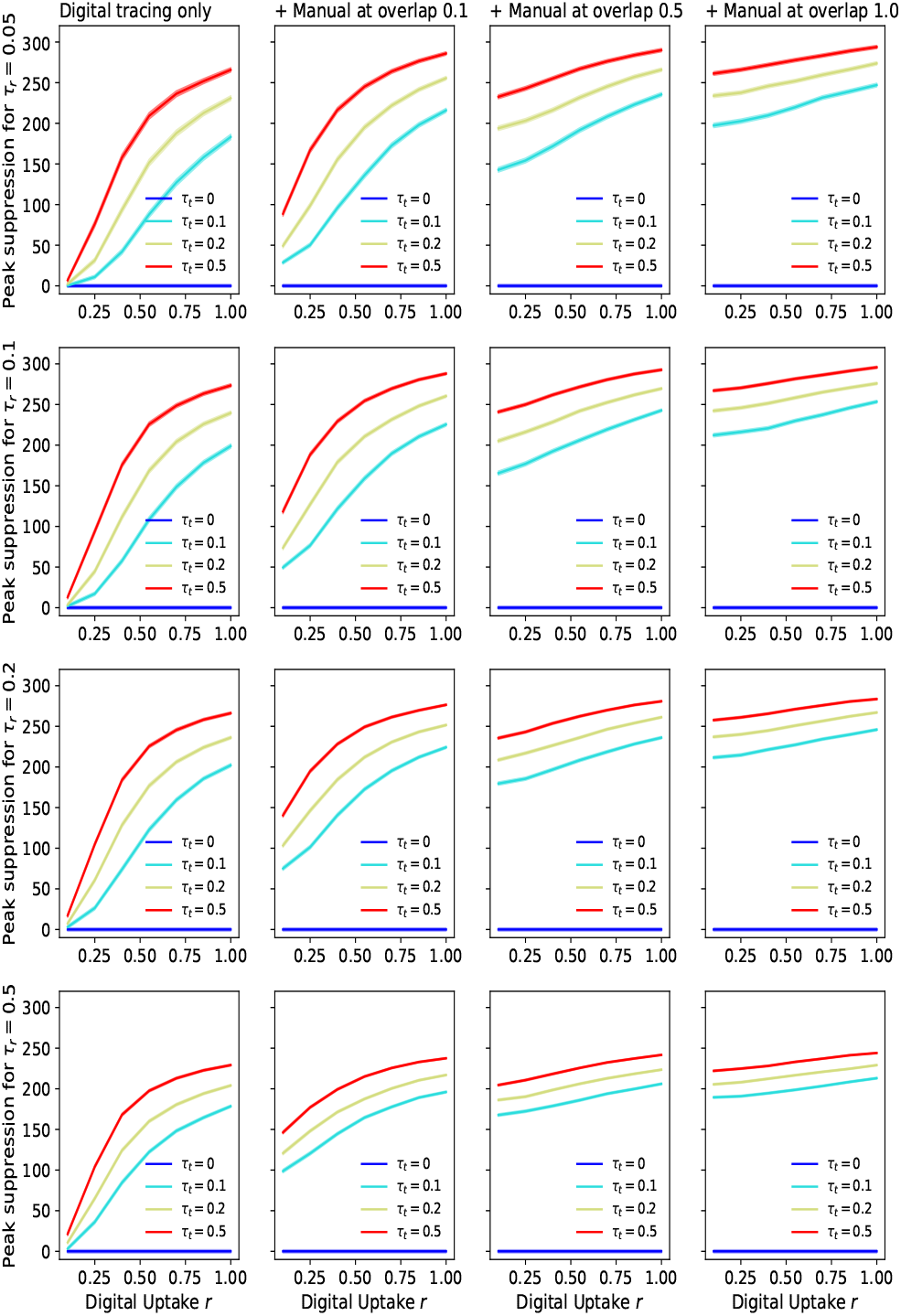
Uptake rate *r* against peak suppression. Suppression is difference in peak to no tracing, i.e. *τ*_*t*_ = 0. The results on the left correspond to a scenario in which only digital tracing was conducted, whereas the next columns represent simulations with a combination of digital tracing on a dual network and manual tracing over a third network with various overlaps (0.1, 0.55, 1). The no-tracing line is colored in blue.

**Figure 10:**
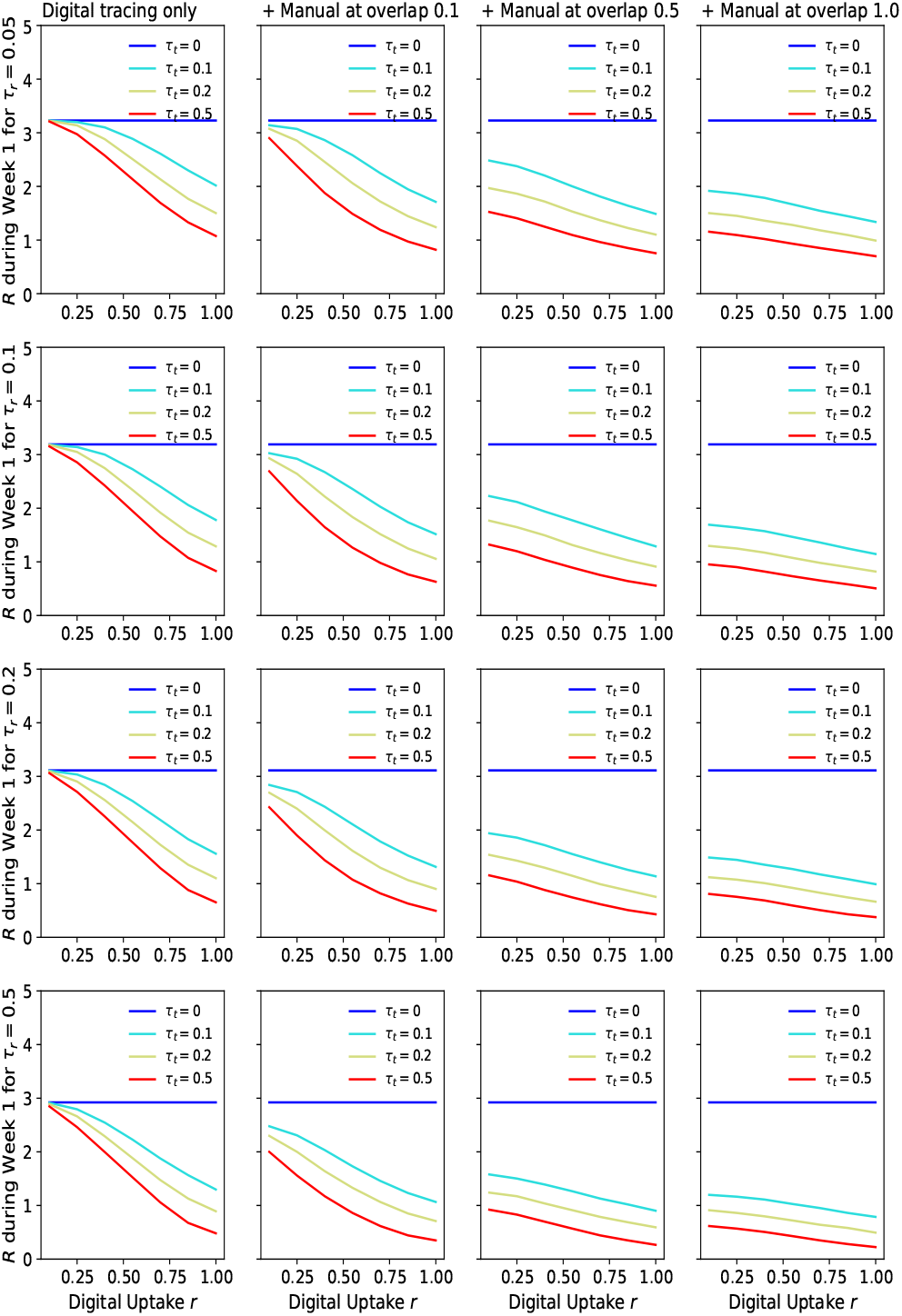
Uptake *r* against the effective reproduction number *R*. Suppression represents the difference to no tracing, i.e. *τ*_*t*_ = 0. The results on the left correspond to a scenario in which only digital tracing was conducted, whereas the next columns represent simulations with a combination of digital tracing on a dual network and manual tracing over a third network with various overlaps (0.1, 0.55, 1). The case with no contact tracing (*τ*_*t*_ = 0) is colored in blue.

Fig 9 shows the degree of peak suppression achieved by utilizing digital and manual tracing solutions when compared to a scenario in which no contact tracing was performed. These results suggest that, as the efficacy of the interview-based process increases (i.e. less contacts get missed), lower and achievable application adoption rates (20-50%) are sufficient to effectively reduce the maximal point of the epidemic. When the tracers are able to “see” the full network of contacts (Γ = 1), varying *r* no longer impacts the spread significantly, as should be expected. In contrast, a very good testing regime (*τ*_*t*_ ≥ 0.2) can partially compensate for an inefficient manual tracing system (Γ = 0.1) within the aforementioned uptake range.

Since the inception of the COVID-19 pandemic, much of the literature on epidemiological modeling and public-health messages alike have scrutinized different nonpharmaceutical interventions in relation with their impact on *R*, the effective reproduction number (Di Domenico et al., 2020; Kajitani and Hatayama, 2020). We estimate our simulations’ *R* value after *t* = 7 days (since *t*_*0*_) from the corresponding exponential growth rate *λ* by applying Eq 1, following the Wallinga and Lipsitch (2007) methodology. The generation time is assumed to form a *Gamma*(*α* = 1.87, *β* = 0.28) distribution (Cereda et al., 2020), denoting with *M* (.) its moment-generating function. To calculate *λ* from the incidence rate *c* (*t*) recorded within the time window [*t*_0_, *t*_0_ + *t*], we use Eq 2 with the infection seed set to *c* (*t*_0_) = 10% (of *N*).

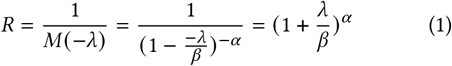

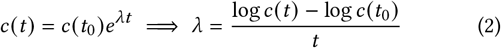

We arrive at an estimate of *R* = 3.20 for minimal interventions (i.e. *τ*_*r*_ = 0.05 and no tracing), value which falls within the confidence interval of the basic reproduction number *R*_0_ [3.09, 3.24] derived in Di Domenico et al. (2020) by applying the next-generation approach (Diekmann et al., 1990) on a model fairly similar to ours. Fig 10 demonstrates that with good testing regimes (*τ*_*r*_ ≥ 0.1 and an efficient manual tracing in place (Γ ≥ 0.5), achievable uptake levels are enough to limit this *R* to a value close to 1. In contrast, digital tracing alone fails to significantly reduce the spread unless both the testing and the adoption rates are very high. Similarly to what can be observed in the peak suppression charts, uptakes play a minor role in the infection progression if tracers are able to track the whole contacts network eventually, yet this scenario is rather unlikely in real life. Interestingly, most of the other trends are faithfully mirrored by the evolution of *R* in the first week of the simulation. This confirms that efficient contact tracing in the early stages of an outbreak is essential for containing such a virus (Shah et al., 2020).

Even though peak suppression remains a good metric for assessing the benefits of public health interventions, policy makers are more often interested in what combinations of these measures can quickly lead to acceptable levels of *R*. In light of this, we plotted the contour lines of the *R* values produced by various degrees of interview-based network overlaps, testing and digital tracing adoption rates (see Fig 11). With an uptake of around 40% in Finland and Ireland, 30% in the UK, or 27% in Germany and Norway at the time of writing ^3^, an effective testing regime (*τ*_*r*_ ≥ 0.2) coupled with an efficient contact tracing rate (*τ*_*t*_ = 0.5) can drive *R* below 1 even when tracers miss up to half the contacts (Γ ≥ 0.5). Should this adoption improve to 50%, the testing rate could be halved and the aforementioned effect would still be obtained. In contrast, a moderate tracing rate only becomes effective if a large-scale testing programme gets deployed (*τ*_*r*_ = 0.5) or bigger uptakes are achieved within a population (*r >* 50%).

**Figure 11:**
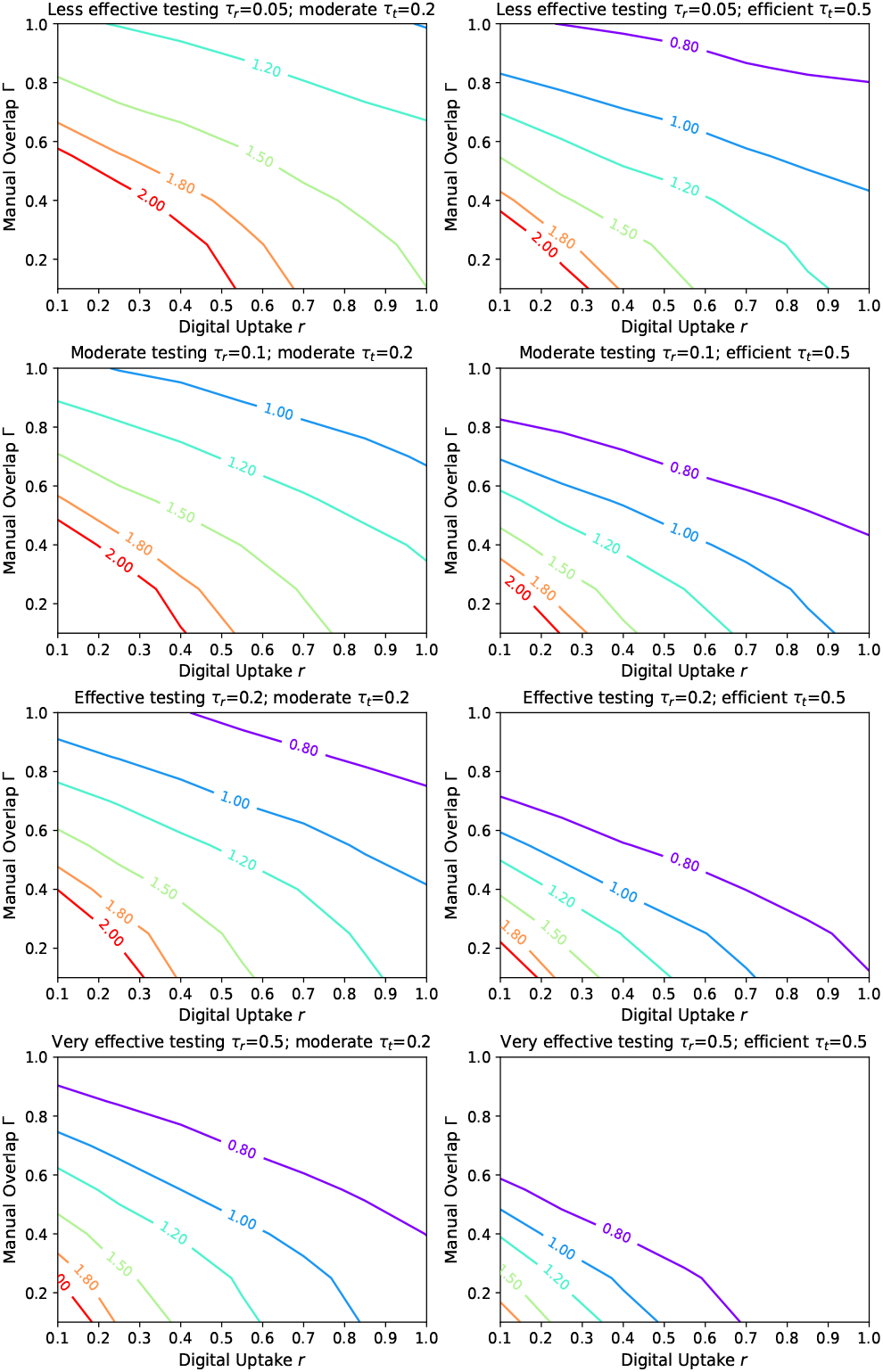
Contour plots of *R* based on the level of manual tracing overlap Γ and digital tracing uptake *r*. Each line represents a different testing level *τ*_*r*_, while the columns correspond to a moderate (left) and an efficient (right) average level of tracing engagement and isolation compliance given by *τ*_*t*_.

## 6 CONCLUSION AND FUTURE WORK

This paper demonstrated how a novel methodology for modelling the effects of different “test and trace” strategies can be applied to study the transmission dynamics of a complex viral outbreak, such as COVID-19. Following a comprehensive analysis of the encapsulated parameters, our SEIR-T model can be used to predict how the SARS-Cov-2 virus would spread through communities where some indication of the interview-based network overlap and/or the digital tracing uptake exists. To facilitate such endeavors, we made our entire codebase open-source (refer to appendix A.1).

The approach we propose can address from a modelling perspective four of the aspects mentioned as open questions in their Cochrane Review by Anglemyer et al. (2020): the combined effects of digital and manual tracing can be studied via the triad network topology, populations with poor access to the internet may be factored in by the degree of overlap Γ, individuals that have privacy concerns or accessibility issues can be represented in the system via the application adoption rate *r*, while the ethical and economical repercussions of balancing false positives and false negatives of tracing can be assessed through the statistics our simulations readily capture. Consequently, our model is powerful enough to answer a large spectrum of research and policy-related questions.

The simulations we conducted show that digital tracing remains effective in reducing the peak of an outbreak, as well as the effective reproduction number *R*, even when the adoption levels are lower. At the same time, a less efficient interview-based process, which misses up to half the contacts, can still contain the spread if coupled with 30-40% application uptakes and large-scale testing regimes. The peak reduction seems to be closely tied to how fast the tracing is conducted, as well as how impactful the public-health messages are in making the community more compliant with the self-isolation recommendation, as soon as more and more of their contacts get traced and isolated (aspects encompassed in the *τ*_*t*_ rate).

We leave for future exploration studying the effects of varying the non-compliance / non-isolation rate, together with the number of initial infections. Looking at such scenarios would allow one to analyze potential reasons for the failing of contact tracing in many countries ^4^, while ensuring a better control over the variability induced by early-stopped simulations.

Next, we envision using mobility datasets to infer more realistic network structures and derive time-dependent estimates of the transmission rate, as previously described by Liu et al. (2020). Other parameters in our model could also be modified according to the epidemiological situation of different countries by fitting them to governmental data reporting on the number of COVID-19 deaths registered within each specific region.

## Data Availability

The data that support the findings of this study were generated using simulations of our open-source model. Most of the raw data we obtained from running these simulations have been deposited on Figshare (link in the Appendix).

https://github.com/andrei-rusu/contact-tracing-model

https://doi.org/10.6084/m9.figshare.14101946

## 7 ACKNOWLEDGEMENTS

We would like to extend our gratitude to Professor Niranjan Mahesan for his lucrative ideas, as well as his continuous effort and support throughout this project. We also acknowledge the use of the IRIDIS High Performance Computing Facility in Southampton.

## A APPENDIX

### A.1 Open-source model and data

The open-source implementation of our model can be consulted at: https://github.com/andrei-rusu/contact-tracing-model.

The statistics our simulations captured can be analyzed in full by following: https://doi.org/10.6084/m9.figshare.14101946.

### A.2 Simulation statistics

Aside from the metrics analyzed in the main text, our model can readily be used to analyze various other statistics about a simulated epidemic: the total number and peak of hospitalization (see Fig S1), total deaths and recoveries, total people that isolated, tracing false positives (Fig S2) and false negatives (Fig S3), the total number of non-compliant nodes, the tracing *efforts*, the incidence and growth rates registered over a variable window size etc.

Both Fig S2 and Fig S3 give an alternative view of the repercussions a country can face if contact tracing is too zealous or too slow. If excessively many people get incorrectly isolated (false positives), the resulting socio-economic burden may significantly disrupt a community. If, on the other hand, tracing is very inefficient, the infectious population (false negatives) will spread the disease uncontrollably, leading to many hospitalizations and deaths. The right balance needs to be struck between these two for a “test and trace” strategy to be deemed successful.

**Figure S1:**
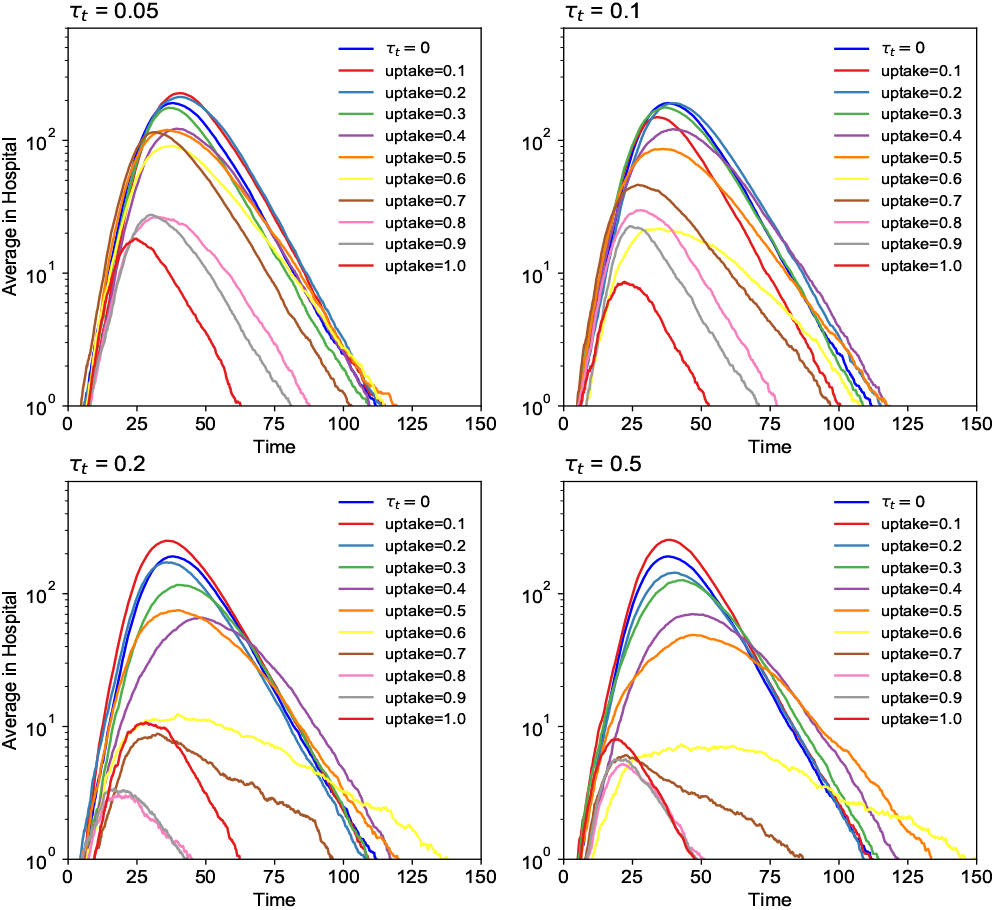
Average number of hospitalizations per unit of time. N=1000, with random graph topology and mean degree *K* = 20. The tracing rate *τ*_*t*_ fixed at 0.1.

**Figure S2:**
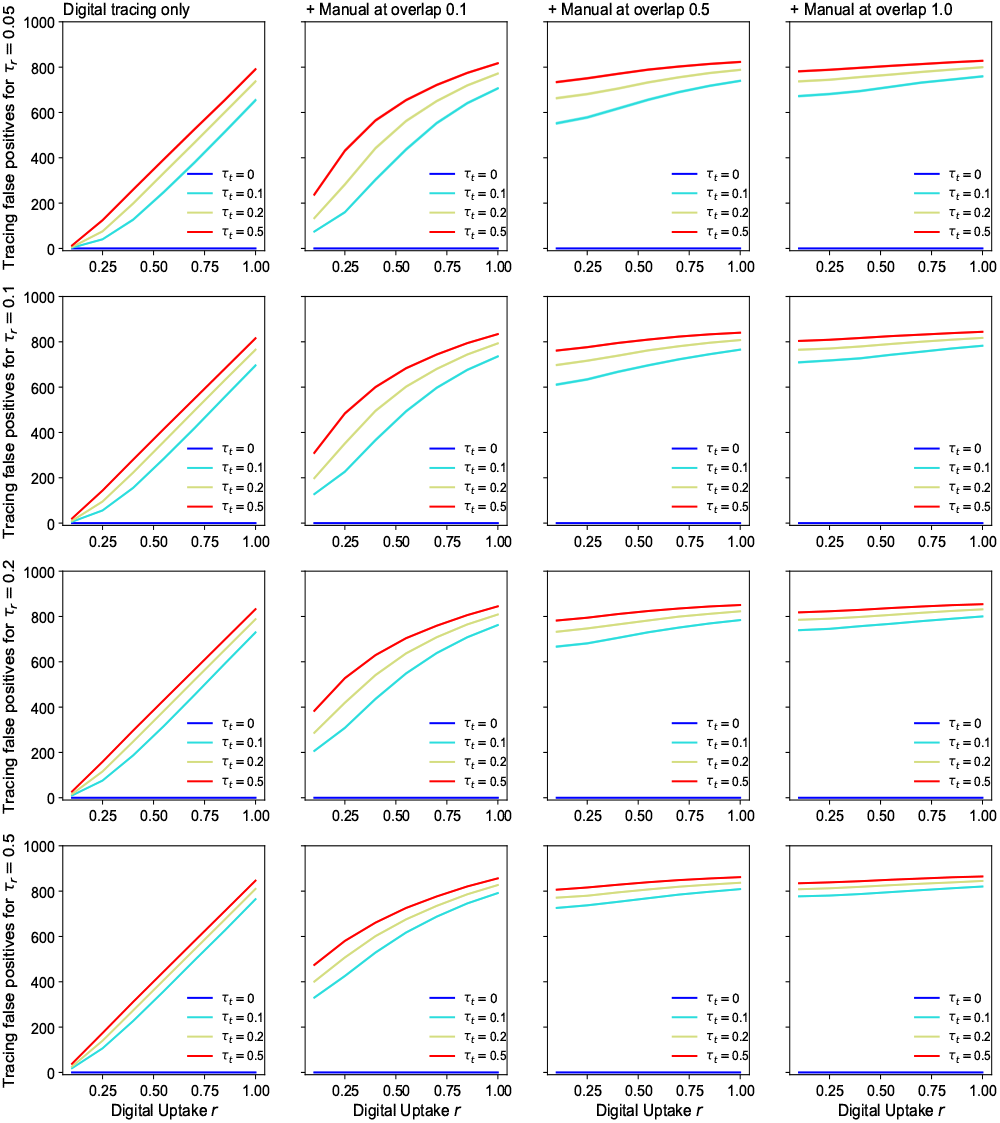
Tracing false positives. The amount of susceptibles being incorrectly traced and isolated. Results here correspond to the last experiment, with *N* = 1000, Holme-Kim graph topology and 10% initial infected.

**Figure S3:**
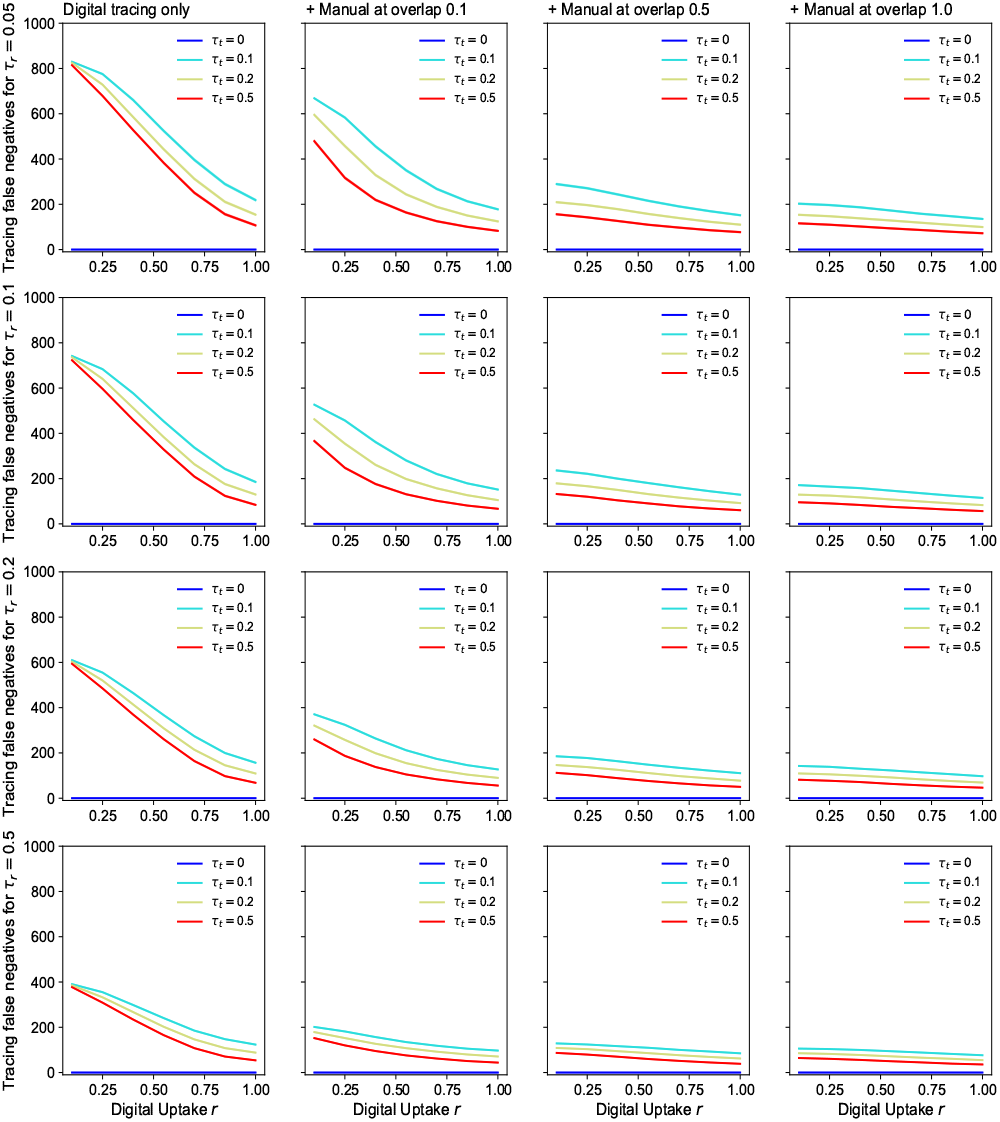
Tracing false negatives. The amount of *infectious* people not traced. Results correspond to the last experiment - *N* = 1000, Holme-Kim graph topology and 10% initial infected.

#### A.3 Further analysis of noteworthy trends

As discussed in the main text, the values ascribed to overlap Γ or uptake *r* (depending on the type of tracing) dictate whether a contact tracing rate *τ*_*t*_ is actually effective. The trends imposed by these quantities on *τ*_*t*_ can be further scrutinized in Fig S4 and Fig S5. At the 0.5 level, both are able to noticeably influence the infection curves obtained by *τ*_*t*_ ≥ 0.04. At the extreme points, the differences in peaks achieved by the same tracing rates become very large and apparent between one another.

When studying the combined effects of manual tracing at different Γ and digital tracing at various *r* on the effective reproduction number *R*, it is often useful to visualize the corresponding three-dimensional trends as a whole. To that end, we plot in Fig S6 the 3D surface of the aforementioned variables belonging to the last experiment.

**Figure S4:**
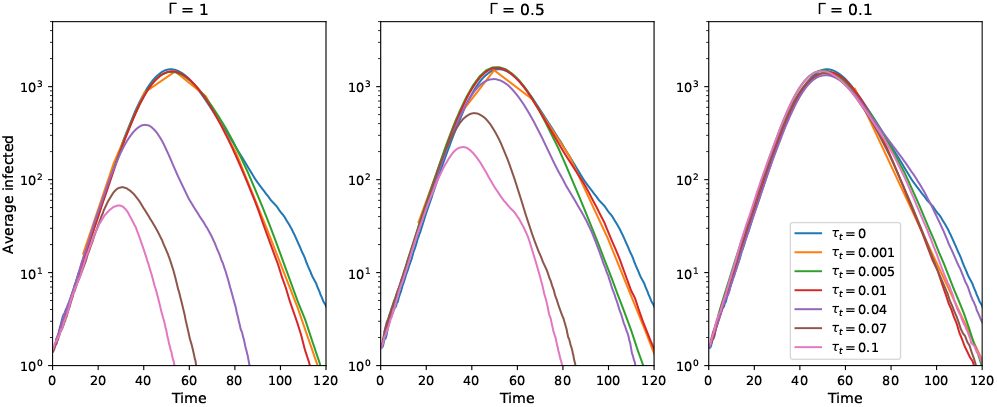
Overlap influencing the efficacy of contact tracing rates. N=10000, random graph topology with mean degree *K* = 10, and *τ*_*r*_ at 0.04.

**Figure S5:**
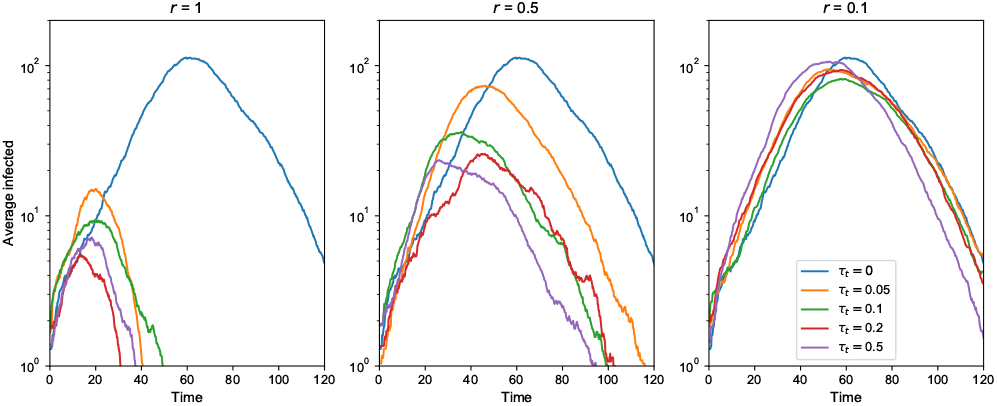
Uptake influencing the efficacy of contact tracing rates. N=1000, random graph topology with mean degree *K* = 10. *τ*_*r*_ fixed at 0.05.

**Figure S6:**
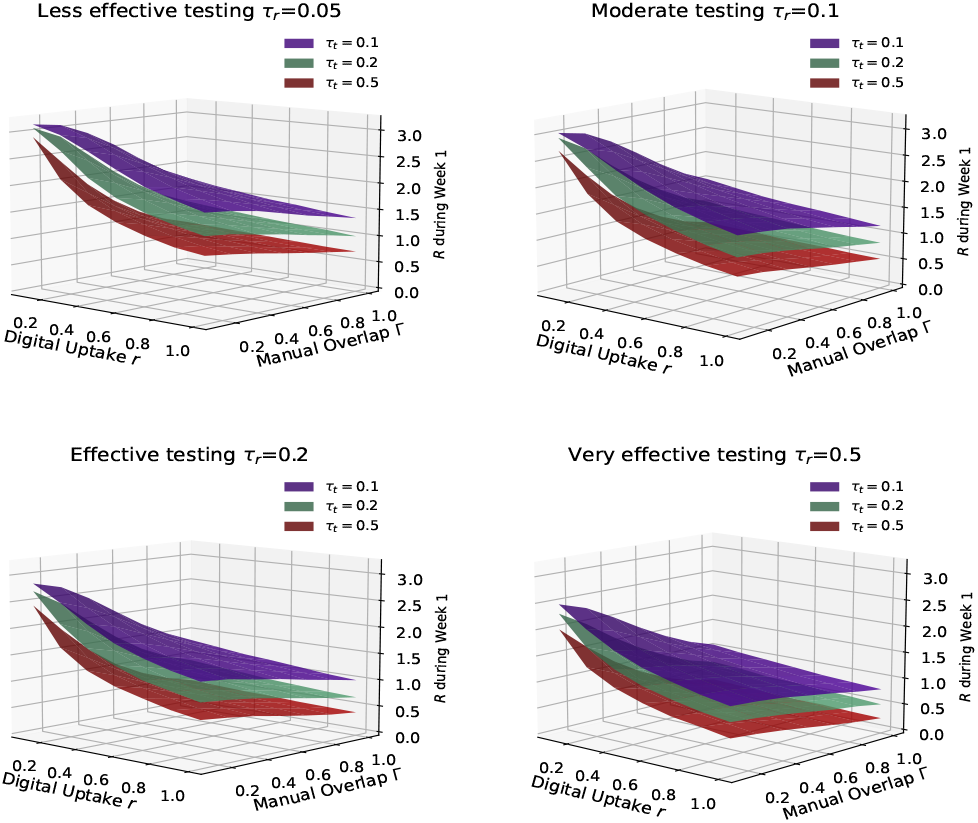
Reproduction number *R* vs. uptake *r* vs. overlap Γ. Results here correspond to the last experiment, with *N* = 1000, Holme-Kim graph topology and 10% initial infected.

## Footnotes

1 https://covid19.who.int

2 https://www.cdc.gov/coronavirus/2019-ncov/php/contact-tracing

3 https://craiedl.ca/gpaw

4 https://www.nature.com/articles/d41586-020-03518-4

